# SOCIODEMOGRAPHIC CHARACTERISTICS AND COVID-19 TESTING RATES: SPATIO-TEMPORAL PATTERNS AND IMPACT OF TEST ACCESSIBILITY IN SWEDEN

**DOI:** 10.1101/2020.12.15.20248247

**Authors:** Beatrice Kennedy, Georgios Varotsis, Ulf Hammar, Diem Nguyen, Germán D. Carrasquilla, Vera van Zoest, Robert S. Kristiansson, Hugo Fitipaldi, Koen F. Dekkers, Meena Daivadanam, Mats Martinell, Jonas Björk, Tove Fall

## Abstract

**Background:** Diagnostic testing is essential for disease surveillance and test-trace-isolate efforts. Here, we aimed to investigate if residential area sociodemographic characteristics and test accessibility were associated with COVID-19 testing rates.

**Methods:** We included information on 421 542 patient-initiated COVID-19 PCR tests from Uppsala County in Sweden from 24 June, 2020 to 9 February, 2022. Using Poisson regression analyses, we investigated whether the Care Need Index (CNI; median 1.0, IQR 0.8, 1.4), a composite measure of sociodemographic factors used in Sweden to allocate primary healthcare resources, was associated with aggregated COVID-19 daily testing rates after adjustments for community transmission. We further assessed if distance to the nearest testing station influenced testing. Lastly, we performed a difference-in-difference analysis of the opening of a testing station targeting a disadvantaged neighbourhood.

**Results:** We observed that CNI, i.e. primary healthcare need, was negatively associated with COVID-19 testing rates in inhabitants aged 5-69 years. More pronounced differences were noted across younger age groups and in Uppsala City, with test rate ratios in children (5-14 years) ranging from 0.56 (95% CI 0.47-0.66) to 0.88 (95% CI 0.81-0.95) across the three pandemic waves. Longer distance to testing station was linked to lower testing rates, foremost in less densely populated areas. Furthermore, the opening of the targeted testing station was associated with increased testing, including twice as high testing rates in individuals aged 70-105, supporting an intervention effect.

**Conclusions:** Ensuring accessible testing across all residential areas constitutes a promising tool to decrease differences and inequalities in testing.

## INTRODUCTION

COVID-19 diagnostic testing has been a cornerstone in the early detection of cluster outbreaks and variants of concern, population-based assessments of COVID-19 vaccine effectiveness, and the evaluation of policy effects (1). Sufficient testing is also essential for successful test-trace-isolate programmes, which helped break the chain of COVID-19 transmission, thereby reducing morbidity (2, 3). However, COVID-19 testing rates have varied considerably even within countries with uniform testing strategies and guidelines, with several studies reporting noting low COVID-19 testing in disadvantaged communities (4, 5, 6, 7, 8, 9, 10). In Sweden, previous studies have reported associations between sociodemographic circumstances and COVID-19 morbidity and mortality (11, 12). Thus, elucidating the sociodemographic determinants of COVID-19 testing within a society may improve testing strategies to successfully reach all inhabitants, thereby reducing adverse COVID-19 outcomes.

In accordance with national guidelines from the Swedish Public Health Agency, free-of-charge population-based COVID-19 PCR diagnostic testing was available in Sweden between June 2020 and February 2022. During this period, each of the 21 Swedish healthcare regions developed their own strategies on how to ensure accessible testing for their populations. In Uppsala County, which has a total population of 386 000 inhabitants and includes Sweden’s fourth most populated city Uppsala, the healthcare council established four main testing stations for patient-initiated tests in June 2020. However, when case notification rates surged in October 2020, additional testing stations were opened in neighbourhoods with low testing rates and indications of high community transmission, and a mobile testing unit was deployed to potential hotspots. The regional testing strategy was subsequently updated to include drop-in testing and the use of self-sampling kits.

In this study, we aimed to investigate the associations between sociodemographic characteristics of residential areas and COVID-19 testing rates in Uppsala County and Uppsala City, from June 2020 to February 2022. Such findings could help explain the previously observed sociodemographic differences in COVID-19 outcomes in Sweden and in neighbouring countries, and provide guidance for future targeted interventions to increase testing for epidemic diseases in disadvantaged communities.

## MATERIAL AND METHODS

### Postal code areas

Sweden is divided into 5-digit postal code areas, of which 533 postal code areas overlap with Uppsala County (Swedish: “Region Uppsala”). We excluded 172 postal code areas that were uninhabited (industrial and/or administrative addresses), 10 that primarily overlapped with neighbouring counties, and 1 for which the Care Need Index had not been calculated by Statistics Sweden (total population <3 residents). After exclusions, we categorized the remaining 350 postal code areas as Uppsala City (postal codes beginning with 75; n=147) and Uppsala County (74 or 81; n=203).

### Care Need Index

We obtained data on the Care Need Index (CNI) for 2020 from Statistics Sweden. CNI is a composite sociodemographic measure calculated for each postal code area in Sweden (see detailed information in Supplementary Material), and is used to allocate primary healthcare resources (13, 14, 15). Higher CNI indicates higher primary healthcare burden. Postal code area CNI constituted our main exposure variable.

### Study period

During the first pandemic wave in March to June 2020, COVID-19 testing in Sweden was only available to healthcare professionals, nursing home residents, and hospital patients (16). From 24 June 2020, updated national guidelines recommended free-of-charge diagnostic PCR testing for all individuals in Sweden aged ≥16 with possible symptoms of COVID-19. Testing in children aged ≥9 was made available on 1 August, 2020, and in children aged ≥5 on 22 February, 2021. Large-scale testing was discontinued on 10 February, 2022. At this time, Sweden was in the throes of the fourth pandemic wave caused by the variant of concern Omicron, and the previous testing strategy was deemed inefficient (17). Our study period was therefore defined as 24 June 2020 to 9 February 2022.

### COVID-19 testing

Information on COVID-19 PCR tests (date of test and test result [positive/negative]) was accessed from the electronic medical records database maintained by Uppsala County Council. After exclusions (see detailed information in Supplementary Material), 434 021 patient-initiated tests were included in the analyses. We did not have access to personal identifiers, and the dataset therefore includes repeated tests from individuals. We aggregated the daily number of tests in sex (women/men) and age groups (5-14, 15-29, 30-49, 50-69, 70-105 years) in each postal code area, and these constituted our main outcome variable.

### COVID-19 case notifications, hospital admissions, and vaccinations

We calculated daily COVID-19 case notification rates per 100 000 inhabitants, per sex, age group, and postal code area across the study period. These case notification rates constituted the main marker of community transmission in our analyses.

We also accessed information on daily COVID-19 hospital admissions, defined as patients with a positive COVID-19 PCR test within 30 days prior to admission or during the hospital stay, per 100 000 inhabitants per postal code area. Information per sex and age group was not available. We used hospital admissions as a secondary marker of community transmission, not affected by testing patterns.

To assess the potential influence of vaccination rates on testing rates across the postal code areas with different CNI, we extracted information on aggregate vaccination coverage in inhabitants aged 15-105 from Uppsala County Council (see detailed information in the Supplementary Material).

### Test accessibility and interventions

During the first part of the study period (24 June to 11 October, 2020), patient-initiated testing was only available at the four main testing stations set up across Uppsala County: in the north, northwest, and east part of the county, and in the centre of Uppsala City (Supplementary Figure 1).

From October 2020 onwards, local health authorities introduced several new measures to encourage testing. In the Uppsala City neighbourhood Gottsunda, local health authorities had noted low testing rates but also high proportion of positive tests and high hospital admission rates. Gottsunda was therefore targeted with a new testing station that opened 12 October 2020. The new test station was highlighted in a press release, which advertised that the staff was multilingual (18).

Several new testing stations were subsequently opened across Uppsala County and Uppsala City, and a mobile testing bus was deployed weekly or biweekly to emerging hotspots (characterized by low testing rates and increasing test positivity). For detailed information on testing availability, drop-in testing, and the use of self-sampling kits, please see the Supplementary Material.

### Statistical analysis

We used multivariable Poisson regression analyses with cluster-robust standard errors (where postal code area represented a cluster) to investigate the association between CNI (exposure) and daily number of tests per sex, age group and postal code area (outcome) across the study period. The natural logarithm (ln) of the population size per sex and age category and postal code was used as an offset in our analysis.

In our main model, we adjusted for date, day of week of test (categorical variable Monday through Sunday; as access to COVID-19 testing varied depending on day of the week), sex and age group, Uppsala County/Uppsala City (binary variable), and daily case notification rates per age and sex group per 100 000 inhabitants per postal code area. Interaction terms were added in the model, with two-, three-, four- and five-fold interactions added between all combinations of date, age group, sex, Uppsala County/Uppsala City, and CNI. Case notification rates and dates were modelled using restricted cubic splines (for details see Supplementary Material).

For each pandemic wave captured by our data, we calculated the highest and lowest test rate ratios (TRRs) per sex and age group in Uppsala County and in Uppsala County, respectively. We defined the start of a pandemic wave as when the 14-day cumulative case notification rates (assessed in Uppsala County and Uppsala City combined) exceeded 500 cases per 100 000, and the end of a pandemic wave when the case notification rates dropped below the same threshold, or when the study period ended, whichever came first. As the first pandemic wave in Sweden occurred from March to June 2020, not captured by our data, we have denoted the three waves in our data as the second (8 November 2020–6 January 2021), the third (20 March–5 May 2021), and the fourth wave (1 January–9 February 2022).

In a sensitivity analysis, we used daily hospital admission rates per 100 000 inhabitants instead of daily case notification rates as marker of community transmission. Hospital admission rates were modelled using restricted cubic splines, with knots placed at -10, 35, 75, and 200 to fit the distribution of the admissions.

To investigate if distance to the main testing station was independently associated with testing, we conducted a separate analysis with distance to main testing station as main exposure, and daily testing rates as outcome, adjusting for all covariates included in the main analysis as well as for CNI. This analysis only encompassed tests from 24 June to 11 October 2020 (when testing was centralized).

To assess any potential intervention effect of the first targeted testing station in the Uppsala neighbourhood Gottsunda, which opened on 12 October 2020, we further performed a difference-in-difference analysis comparing testing rates in Gottsunda with the Uppsala City neighbourhood Sävja between 12 July 2020 and 12 January 2021 (see additional details in the Supplementary Material). Sävja was matched to Gottsunda as it also represented a disadvantaged neighbourhood located more than 5 kilometers from the City centre and the Uppsala City main testing station. However, Sävja was, in contrast to other similar neighbourhoods, not targeted by any testing efforts by local healthcare authorities in October to December 2020.

Finally, we generated graphs of model-based testing rates per 100,000 per sex, age category and Uppsala city/county by setting day of the week to Wednesday and daily case notification rates (or, in the sensitivity analysis, hospital admissions rates) to zero. We used Uppsala County and Uppsala City-specific CNI 10^th^ and 90^th^ percentiles. Similar graphs were constructed for model-based testing rates with distance to main testing station as the exposure, as well as for the difference-in-difference analysis on Gottsunda/Sävja.

All analyses were conducted using Stata 15 (10). Maps and plots were produced using R software (v. 4.1.3, March 2022)(19). The ‘tmap’ package (v. 3.3.3)(20), ‘ggplot2’ package (v. 3.3.6)(21), the ‘ggpattern’ (v. 0.4.2)(22), and the fontawesome package (v. 0.4.0)(23) were used to produce the maps and plots.

### Ethical statement

This study was approved by the Swedish Ethical Review Authority (DNR 2020-04210 and DNR 2021-01915).

## RESULTS

### Baseline characteristics

Baseline characteristics of the 350 postal code areas in Uppsala County and Uppsala City are presented in Figure 1 and Supplementary Table 1. CNI across all postal code areas ranged from 0.0 to 3.2 (Supplementary Figure 2), with a lower median noted in Uppsala County (0.8, IQR: 0.7, 1.1) than in Uppsala City (1.1, IQR: 0.8, 1.6;). Compared to Uppsala County, the postal code areas in Uppsala City had lower median age, higher proportions of women and of inhabitants born in areas outside of the European Union. Furthermore, we observed that CNI in both Uppsala County and Uppsala City were strongly positively correlated with proportion of inhabitants who had compulsory education only or were born in areas outside of the European Union (Supplementary Figure 3). From 24 June to 11 October 2020, the distance to the nearest main testing station was longer for postal code areas in Uppsala County than in Uppsala City.

**Figure 1.**
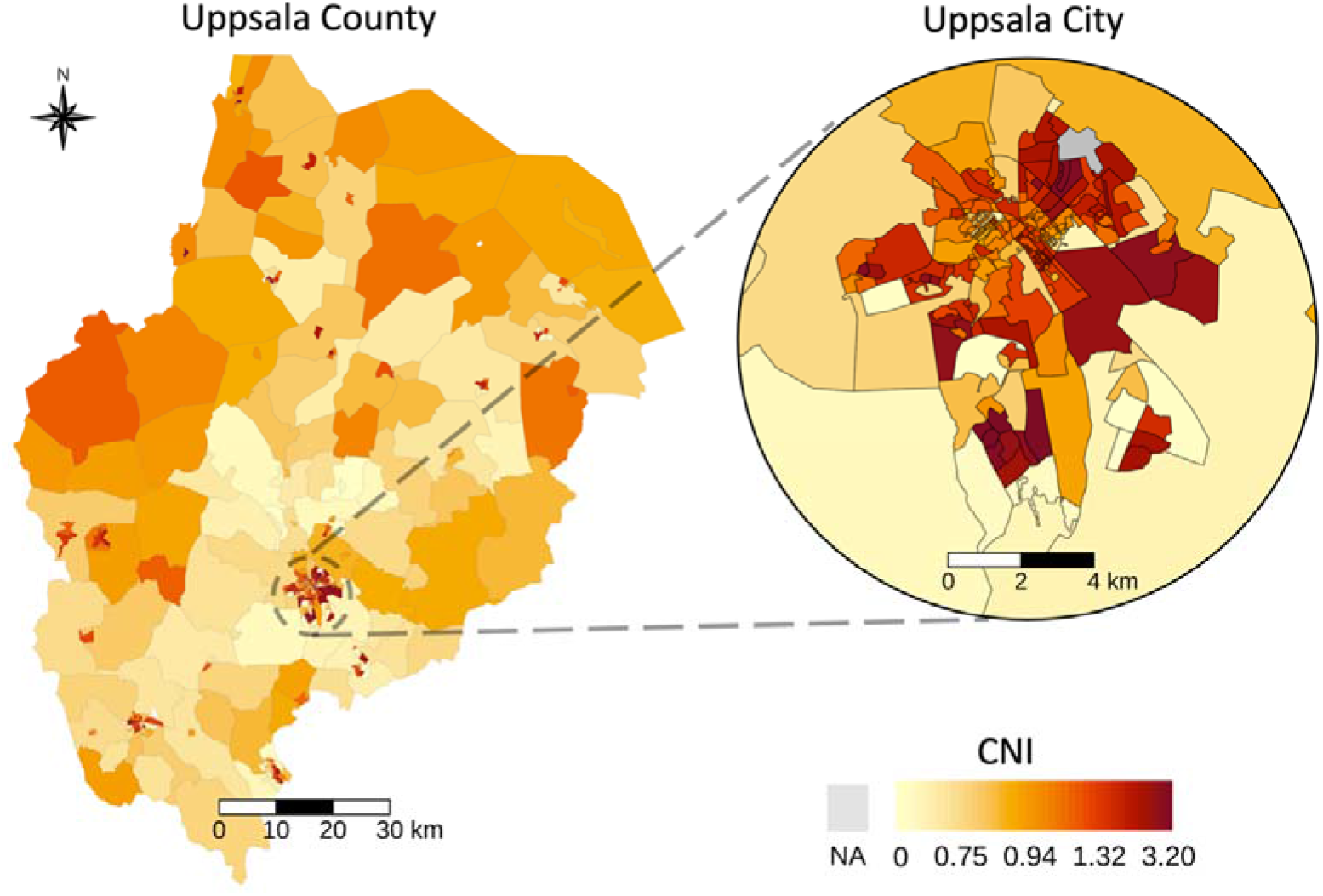
Geographical distribution of Care Need Index (CNI) across postal code areas in Uppsala County and Uppsala City. Higher CNI indicates higher primary health care burden. The colour legend changes non-linearly to facilitate visualisation of the skewed distribution.

The two neighbourhoods Gottsunda and Sävja in Uppsala City both had higher CNI than the Uppsala City average (Supplementary Table 2). Compared to Sävja, Gottsunda had a higher proportion of inhabitants born in areas outside of the European Union, unemployed, or with compulsory education only.

### COVID-19 testing, case notification and hospital admission rates

Testing and case notifications rates demonstrated similar patterns in inhabitants from Uppsala County and Uppsala City across the study period, with the three peaks of case notifications coinciding with the three peaks of hospital admissions (Supplementary Figure 4). The highest testing rates were observed in Uppsala City (7-day rolling average of >400 tests per 100 000) in April and December 2021, the lowest in June to August 2020 and 2021. Inhabitants aged 70-105 had the overall lowest testing rates across age groups (Supplementary Figure 5). Testing rates were higher in women aged 15-69 than in men, with the largest difference noted in women aged 30-49. Testing rates did not differ notably by sex in children aged 5-14 or in inhabitants aged 70-105.

### COVID-19 vaccination coverage

The population-weighted median cumulative vaccination coverage in inhabitants aged 15-105 years reached 88.3 (IQR 85.1, 90.4) in Uppsala County and 88.5 (IQR 83.3, 90.4) in Uppsala City on 9 February, 2022. Across CNI quartiles, we observed an inverse association between CNI and vaccination coverage (Supplementary Figure 6).

### CNI was associated with COVID-19 testing rates

Throughout the study period, we observed a negative association between CNI and testing rates in several age groups (aged 15-69) in both women and men (Figure 2 and Figure 3). These patterns were more pronounced in Uppsala City than in Uppsala County. Further, CNI was consistently negatively associated with testing rates in children aged 5-14 across all three pandemic waves in both Uppsala City and Uppsala County (Supplementary Table 3), with test rate ratios (TRRs) ranging from 0.56 (95% CI 0.47-0.66) to 0.88 (95% CI 0.81-0.95).

**Figure 2.**
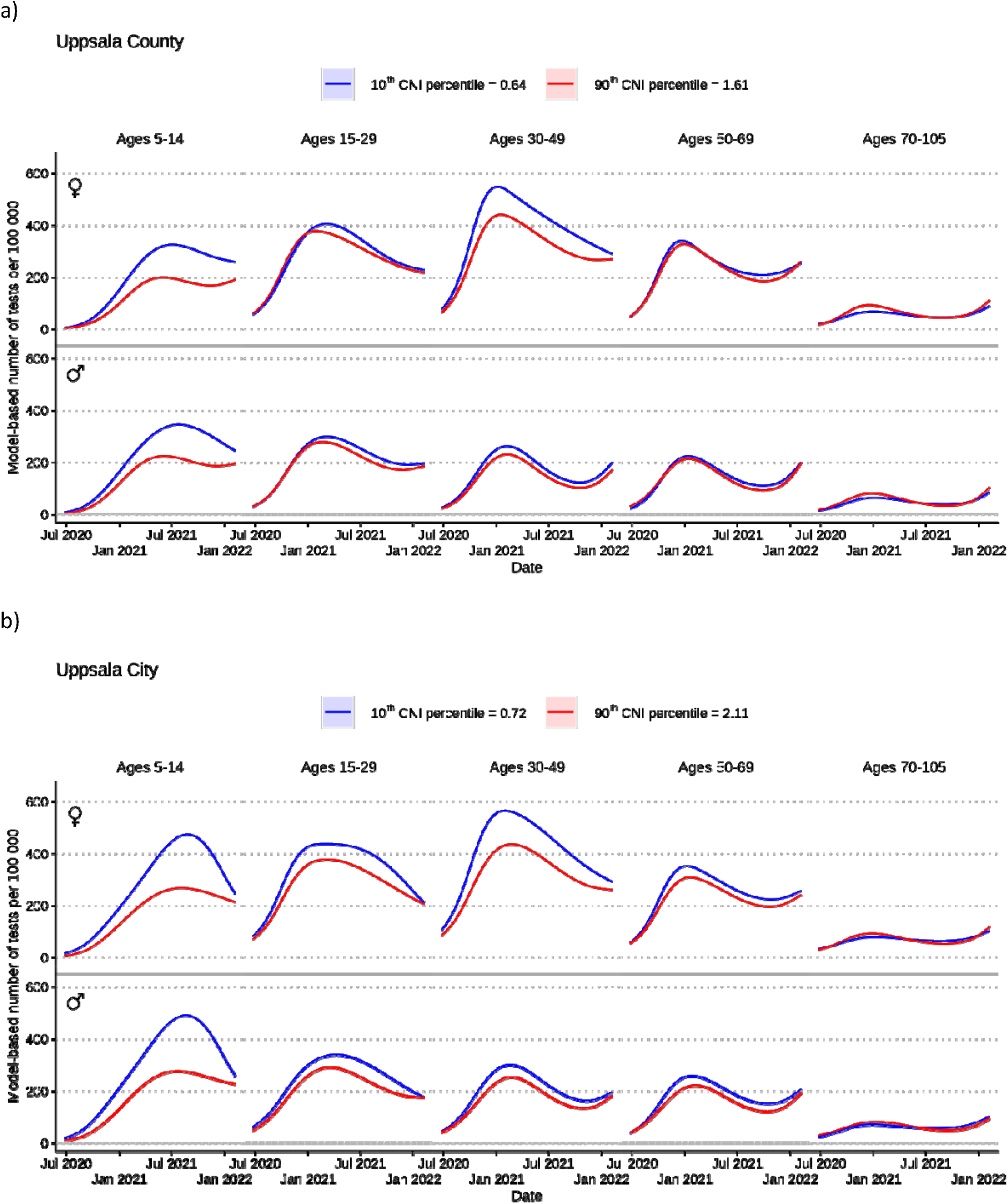
Model-based testing rates for COVID-19 per sex and age group per 100 000 inhabitants in Uppsala County (not including Uppsala City) (a) and Uppsala City (b) across the study period (24 June 2020–9 February 2022), presented by 10^th^ and 90^th^ postal code area Care Need Index (CNI) percentiles. Higher CNI indicates higher primary health care burden.

**Figure 3.**
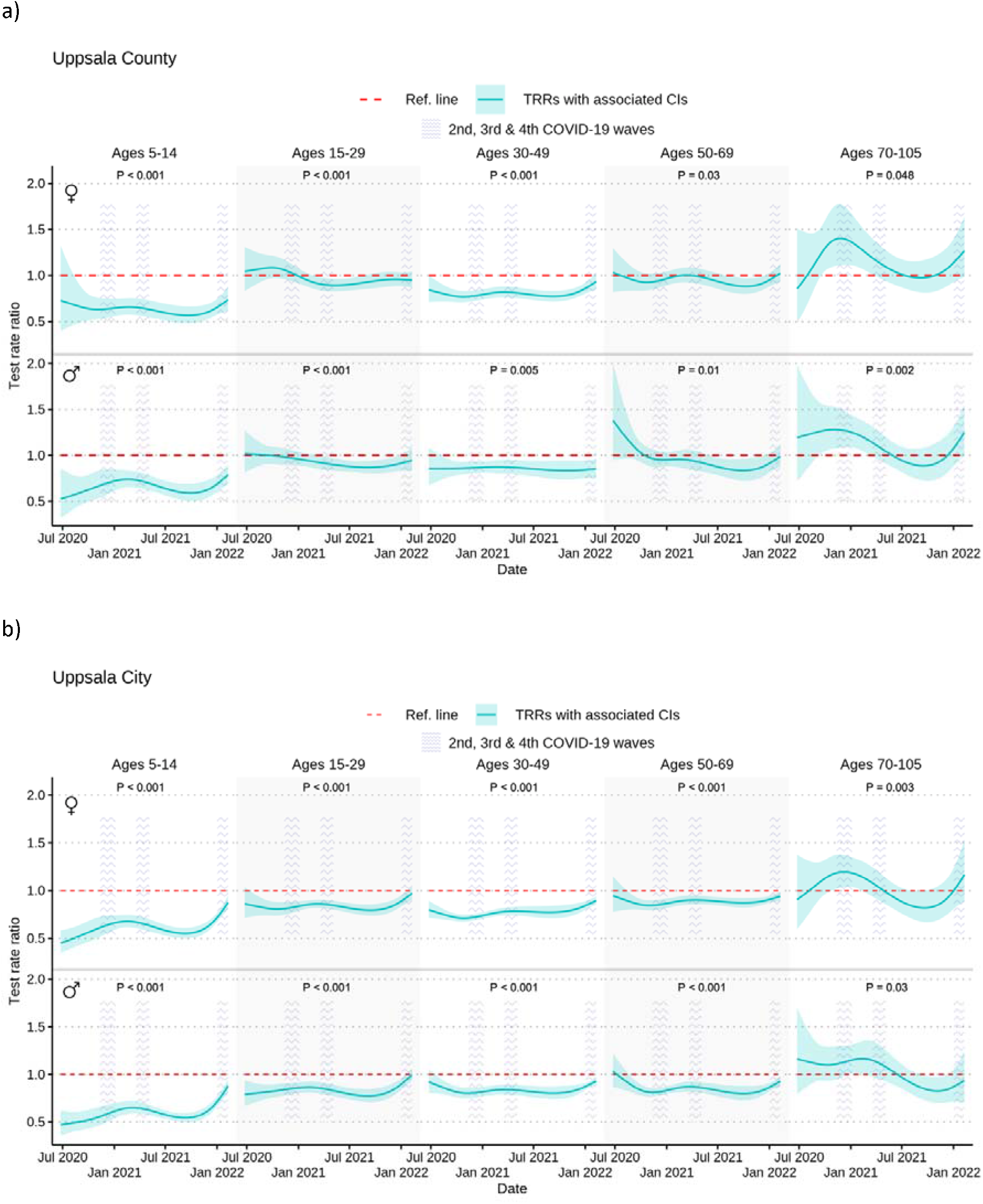
COVID-19 test rate ratios (TRRs) and 95% confidence intervals (CIs) by postal code area Care Need Index (CNI) per sex and age group per 100 000 inhabitants in Uppsala County (not including Uppsala City) (a) and Uppsala City (b) across the study period (24 June 2020–9 February 2022) and the three pandemic waves captured by our study.

In contrast, in women aged 70-105 in both Uppsala County and Uppsala City, and in men aged 70-105 in Uppsala County, CNI was positively associated with testing rates during the second pandemic wave (12 November 2020 to 5 January 2021), with TRRs ranging from 1.18 (95% CI 1.04-1.35) to 1.40 (95% CI 1.11, 1.77). We could not detect any such associations in these age groups during the subsequent waves.

Our sensitivity analysis, where we employed daily hospital admissions instead of daily case notifications as a marker of community transmission, yielded similar results as the main model (Supplementary Figure 7, Supplementary Table 4).

### Distance to testing station and the intervention effect

We found that longer distance to testing stations was independently associated with lower testing rates, after adjusting for CNI, foremost in younger age groups in Uppsala County (Supplementary Figure 8, Supplementary Table 5), and in children aged 9-14 years in Uppsala City.

We further observed that the opening of a targeted testing station in Gottsunda on 12 October 2020 coincided with increased testing rates in Gottsunda, as compared to the non-targeted neighbourhood Sävja (Figure 4). The difference-in-difference analysis indicated an overall intervention effect (omnibus p-value <0.001). We observed twice as high testing rates in both women and men in the oldest age groups (70-105 years), and a large but transient difference in testing rates in girls aged 9-14. The patterns in the other age groups were less pronounced.

**Figure 4.**
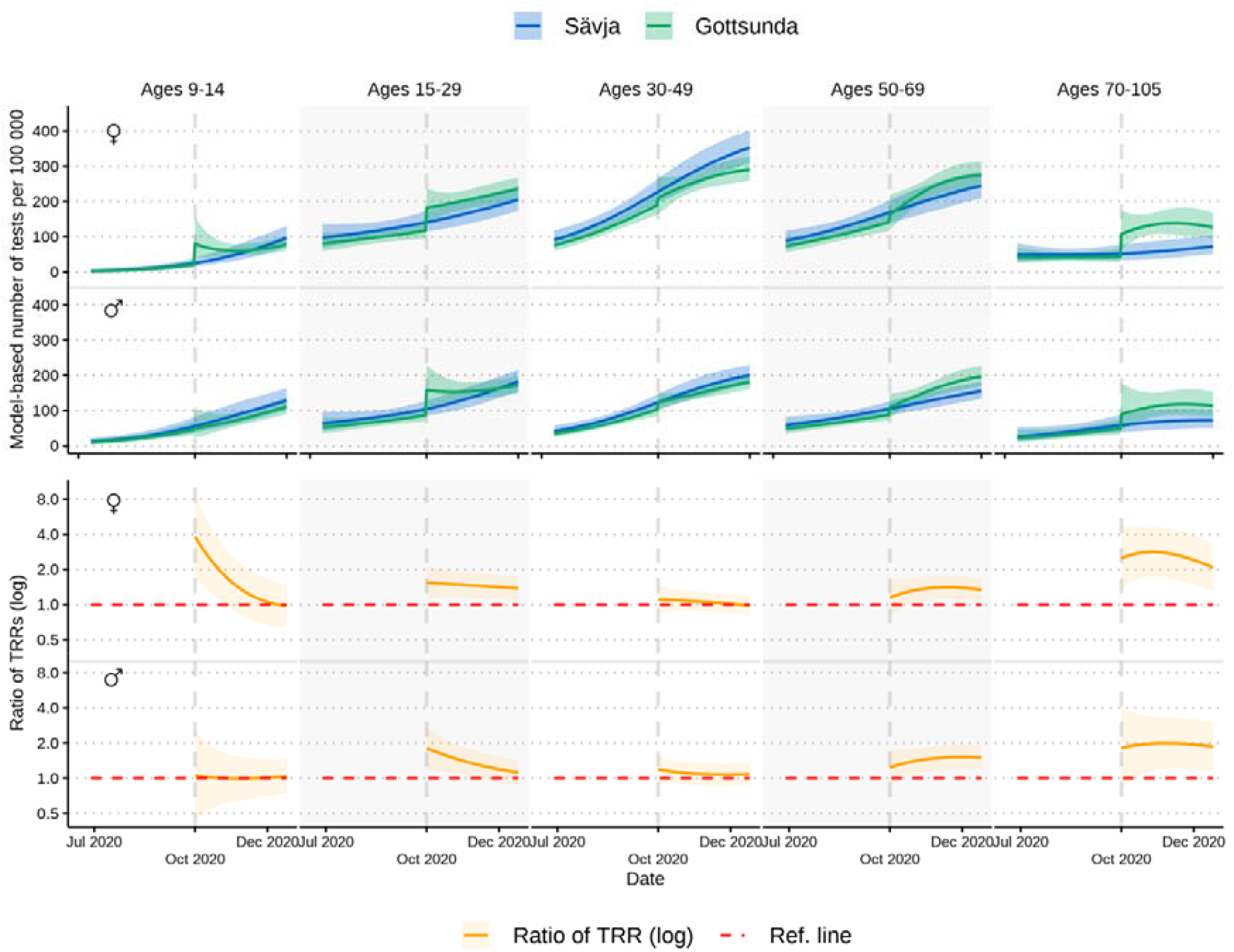
COVID-19 testing in the Uppsala City neighbourhoods Gottsunda and Sävja 12 July 2020–12 January 2021, with Gottsunda targeted by local health authorities with a local testing station, opened on 12 October 2020. Top panels depict model-based testing rates per sex and age group per 100 000 inhabitants in Gottsunda and Sävja. Bottom panels depict results from difference-in-difference analysis (ratio of test rate ratios [TRRs]) comparing Gottsunda and Sävja per sex and age group. Higher ratio of TRRs indicates relative increase of testing in Gottsunda (omnibus p-value <0.001).

## DISCUSSION

This study aimed to investigate the association between residential area sociodemographic characteristics and COVID-19 testing rates in Sweden. We found that postal code area CNI was negatively associated with testing rates in both adult and child inhabitants across the study period, which encompassed the three separate pandemic waves. Additionally, longer distance to the nearest testing station was associated with lower testing rates, more pronounced in Uppsala County that included less densely populated areas. Lastly, we noted that the opening of a new test station in a disadvantaged neighbourhood with high community transmission rates was linked to an increase in testing.

Previous studies on COVID-19 outcomes have used different definitions of neighbourhood sociodemographic characteristics (4, 5, 6, 7, 8, 9, 10). In this study, we used the Swedish composite measure CNI, which incorporates seven weighted sociodemographic area variables. CNI is correlated to other established area deprivation scores such as the Townsend Deprivation Index (15), and higher CNI has previously been linked to poorer health in both adults and children (14, 24, 25). More importantly, CNI is currently employed by the Swedish health authorities to allocate primary healthcare resources, and thereby constitute a readily available tool for allocating and evaluating COVID-19 test efforts.

Our findings align with an earlier report from the Skåne healthcare region in Sweden using data from June 2020 to April 2021, which also linked residential area sociodemographic factors to lower testing rates (26). Our observations further correspond with reports from the United States and the United Kingdom (5, 6, 7, 8, 9), as well as with Swiss studies comprising data on COVID-19 testing rates, hospitalizations, and deaths in 2020–2021 (4, 10). A study investigating COVID-19 mortality in Stockholm, Sweden, found that many migrant groups were particularly vulnerable during the first pandemic wave, but that part of the excess risk could be attributed to demographic factors such as neighbourhood population density (12). The importance of neighbourhood characteristics, independent of individual-level risk factors, was also highlighted in a later report from the Stockholm County Council on COVID-19 mortality, indicating an interplay between individual risk factors and residential area circumstances (11). As our study does not include individual-level data, we cannot distinguish the influence of residential- and individual-level factors on testing, and both likely contributed to our findings. At the end of the study period, we observed that CNI was also inversely associated with vaccination coverage.

Interestingly, we observed that during the second pandemic wave between November 2020 and January 2021, testing rates were higher in inhabitants aged 70-105 years residing in areas with higher CNI than in those residing in areas with lower CNI, a pattern not noted during subsequent pandemic waves. It is possible that the higher testing rates were attributable to higher community transmission not captured by our models, or to a higher prevalence of health risk factors that entailed a lower threshold for testing at that time. However, it is also possible that older age groups residing in more affluent neighbourhoods could more easily adhere to the 2020 national COVID-19 recommendations of physical distancing for individuals aged ≥70. This would correspond to a Norwegian COVID-19 study that reported a positive association between individual-level income and education level and adherence to non-pharmaceutical interventions such as physical distancing and avoidance of public transportation in 2020 (27).

We observed that longer distances to the nearest testing station were associated with lower testing rates in younger age groups, even after adjusting for CNI. This finding is consistent with previous studies that indicated worse health outcomes and lower adherence to health prevention and screening programmes among individuals living farther away from healthcare facilities, compared to those who lived closer (28, 29). For future successful COVID-19 test efforts, the influence of longer distances to test facilities may need to be carefully considered, especially as public transport is commonly discouraged for individuals with COVID-19-related symptoms.

We leveraged a difference-in-difference-analysis, a quasi-experimental design that can employ longitudinal data to estimate causal effects of interventions (30), to evaluate the targeting of Gottsunda with a testing station. The opening was accompanied by an information campaign and was highlighted in local media outlets. Our results supported an intervention effect, and we noted a large increase in testing in the oldest age groups that are also the most vulnerable to adverse COVID-19 outcomes. Our findings emphasize that neighbourhoods with low testing rates should be subject to testing interventions tailored to that specific community.

### Strengths and limitations

Strengths of the study include the use of detailed data extracted from population and health registers on postal code area sociodemographic characteristics and COVID-19 testing, case notification and hospital admission rates. All PCR tests conducted in Uppsala County and City were administered and analysed by the Uppsala County Council, ensuring that our study comprises all tests from a Swedish county, which had uniform testing recommendations and test booking procedures across the study period. Furthermore, our data captured three separate pandemic waves from 2020 to 2022. Some potential limitations apply. Firstly, the use of aggregate data on sociodemographic circumstances and on testing, also including repeated tests, limits the application of our findings to individuals, and thus, contextual and individual effects cannot be disentangled. Secondly, even though we adjusted for case notification rates, and for hospital admission rates in a sensitivity analysis, we cannot exclude the possibility of residual confounding by differences in community transmission not fully captured by our models. Thirdly, we cannot ascertain that our findings are generalizable to other healthcare regions in Sweden with different testing strategies, population compositions and geospatial structures. For example, the distance to the nearest testing station may be even more influential in the eight northernmost healthcare regions in Sweden, each occupying more than twice the geographical area of Uppsala County and with markedly lower population density. Lastly, our results may not apply to other countries with different testing strategies that relied more on lateral flow tests and/or preventive screening of asymptomatic individuals.

### Conclusion

We observed sociodemographic disparities in COVID-19 testing rates across residential areas in a Swedish county with uniform guidelines on testing, but also that a targeted test intervention was associated with higher testing rates. Our findings highlight the importance of considering local context and community-specific factors when designing public health interventions.

## Data Availability

Restrictions apply to the availability of the testing and vaccination data, which were used under license and ethical approval. The data are however available from the authors upon reasonable request and with written permission from the Ethical Review Authority in Sweden. Information on postal code areas included in analyses and the code used in the project are available on Github (https://doi.org/10.5281/zenodo.7919372).

## ACKNOWLEDGEMENTS

The computations and data handling were enabled by resources from project sens2020559 provided by the National Academic Infrastructure for Supercomputing in Sweden (NAISS) and Swedish National Infrastructure for Computing (SNIC) at Uppsala Multidisciplinary Center for Advanced Computational Science (UPPMAX), partially funded by the Swedish Research Council through grant agreements no. 2022-06725 and no. 2018-05973.

## AUTHOR CONTRIBUTIONS

BK, MM, UH, JB and TF conceived and designed the study. UH performed the statistical analyses, BK wrote the first draft, and GV designed the maps and figures. BK, MM, UH, JB, TF, DN, GDC, VvZ, RSK, HF, KFD, MD together interpreted the findings, and reviewed, edited, and approved the final article. TF is the principal investigator, has had full access to all the data in the study, and accepts final responsibility for the decision to submit for publication.

## Key points

- Free-of-charge COVID-19 PCR diagnostic testing was available to the public in Uppsala County and Uppsala City, Sweden, from 24 June 2020 to 9 February 2022, in accordance with the national guidelines from the Swedish Public Health Agency.
- Across the study period, which included three separate pandemic waves, we observed lower COVID-19 testing rates in residential areas with higher Care Need Index, a composite measure of area sociodemographic factors used to allocate primary healthcare resources in Sweden.
- COVID-19 testing was initially centralized at four main testing stations across Uppsala County and Uppsala City, and, during this period, longer distance to the nearest testing station was associated with lower testing rates in younger inhabitants and in less densely populated areas.
- The opening of a local testing station in a disadvantaged neighbourhood was associated with a subsequent increase in testing rates noted across all ages, and with twice as high testing rates in inhabitants aged 70-105.
- Ensuring accessible COVID-19 testing across all residential areas may constitute an effective tool for decreasing differences in testing rates.

