## Supplementary Material for "SOCIODEMOGRAPHIC CHARACTERISTICS AND COVID-19 TESTING RATES: SPATIO-TEMPORAL PATTERNS AND IMPACT OF TEST ACCESSIBILITY IN SWEDEN"

**The Care Need Index and choropleth maps**

The composite measure Care Need Index (CNI) is computed using seven demographic and socioeconomic variables, i.e. proportion of inhabitants in a postal code area who are 1) <5 years old, 2) born in eastern Europe (outside the European Union), Asia, Africa, or South America, 3) >65 years and reside in single-person households, 4) single parents with children <18 years old, 5) >1 year old and have moved into the postal code area within the previous calendar year, 6) 25-64 years old with low educational attainment (≤9 years of schooling, equivalent to compulsory education only in Sweden), 7) 16-64 years old and unemployed or enrolled in a labour market programme (for additional information, see Supplementary Material). The weight of each variable depends on the influence on primary healthcare workload, as determined by individual scoring by a panel of primary healthcare physicians. (1-3)

To calculate Care Need Index (CNI) for each postal code area in Sweden in 2020, Statistics Sweden performed the following sequential steps: To assess CNI per person, the sum of all seven sociodemographic variables, calculated as the number of individuals per variable multiplied by variable-specific weight, was divided by the total number of inhabitants per postal code area. Then, the CNI per person was divided by the CNI per person for the median CNI postal code area, which was 2.13.

We created choropleth maps of CNI per postal code area across Uppsala County and Uppsala City. We generated the geographic features by using administrative shape-files derived from an external company, Postnummerservice Norden AB, containing detailed information on five-digit postal code area boundaries. The resulting geospatial polygons were coloured with light-to-dark sequential schemes to represent the CNI distribution; darker colours represent higher CNI. The colour legend changes non-linearly to facilitate visualisation of the skewed distribution.

**COVID-19 tests**

In total, 625 786 PCR diagnostic tests were performed during the study period in individuals with a registered home address in any of the 350 postal code areas included in this study. We excluded tests from non-symptomatic individuals performed as part of the screening and tracing efforts within healthcare and/or elderly care (N=107 423), tests that were requested by a physician and therefore not patient-initiated (N=96 688), and tests performed in individuals aged <5 or >105 (N=40). We further excluded tests in children aged <9 years before 1 August 2020 (N=25) and in children aged <5 years before 22 February 2021 (N=68) as these tests did not conform to testing guidelines. The resulting dataset consisted of 434 021 tests included in our analyses.

**COVID-19 testing availability**

During the first part of the study period (24 June–11 October 2020), patient-initiated COVID-19 PCR tests were conducted at for the four main testing stations set up across Uppsala County - in the north, east, and northwest part of the county, and in the centre of Uppsala City. Tests were pre-booked through the national online healthcare platform Healthcare Guide 1177.se (hereafter, 1177). All time slots for tests were made available in three-day rolling increments. The online booking required access to electronic identification by BankID, a national identification certificate available for tablets, smart phones, and computers. Individuals who did not have a Swedish personal identification number and/or a BankID could book a test by calling the number provided after initial contact with the national 1177 or a primary care centre. Online booking was only available in Swedish, while phone booking was available in both Swedish and English, and interpreters for other languages were available when necessary. Test for children aged 5-12 were pre-booked by their legal guardians. For children aged 13-16, the child could pre-book their own test if they had a FrejaID (also a national identification certificate), if not, their parents booked tests by phone. All testing was performed by medical personnel who conducted nasopharyngeal and oropharyngeal swabs. No self-test kits were distributed and no drop-in testing was available during this time. Asymptomatic individuals were not declined testing.

A dedicated testing station was established in Gottsunda on 12 October 2020, and was the first additional testing station opened by the local health authorities. Between October 2020 and April 2021, six more testing stations were opened across Uppsala County (in the municipalities of Håbo, Knivsta, Heby and Älvkarleby) and Uppsala City (in the neighbourhoods Löten and Stenhagen).The testing capacity at the local primary care unit in the municipality of Knivsta was also expanded. Between 16 November 2020 and 12 February 2021, a bus serving as a mobile test unit was deployed weekly or biweekly to six emerging hotspots characterized by low testing rates, but high test positivity, within Uppsala City and across Uppsala County. During the winter of 2020–2021, drop-in testing was also initiated at several testing stations in Uppsala County and Uppsala City.

On November 29, 2021, the regional testing strategy transitioned from one focused on health-care administered tests to one focused on self-test PCR kits, which could be collected at the testing stations without prior appointment, registered through 1177 and then dropped of any testing station for analyses. However, due to extreme high demands for these self-test kits in Uppsala City, collection of a self-test kit required pre-booking at 1177 from 15 December 2021. PCR testing of the general public was discontinued from 10 February 2022

During the entire study period, the Uppsala County Council initiated several communications and media campaigns related to COVID-19 testing, including information about COVID-19 symptoms and the importance of social distancing. The opening of new test stations and deployment of the mobile test units were announced during news conferences.

**Distance to main testing station**

We determined driving distance from each postal code area to the nearest main test station (July 24 to October 11, 2020) by Google Maps Distance Matrix API using the R package gmapsdistance in R version 4.0.32. The Distance Matrix created a polygon of each postal code area, and the centre of the polygon was defined as the postal code location. The location of each of the main test stations was defined by the street address.

**COVID-19 vaccinations**

The COVID-19 vaccination programme was initiated in Uppsala County and Uppsala City in January 2021. At the end of our study period, all inhabitants ≥12 years had been invited for vaccination, as well as small number of children aged 5-11 with severe chronic medical conditions (4, 5). We obtained aggregate data on vaccine coverage from the Uppsala County Council, and calculated population-weighted time-updated cumulative vaccine coverage (defined as ≥2 doses) in inhabitants 15-105 years across postal code areas by CNI quartiles. In accordance with the legal regulations of healthcare in Sweden, residents of Uppsala County and Uppsala City were eligible to receive COVID-19 vaccinations in other parts of Sweden. However, we did not have access to vaccination data from other parts of the country in our study.

**Restricted cubic splines for case notification rates and dates**

Knots for splines of case notification rates per 100 000 were placed at -10, 5000, 20 000 and 40 000, Knots were chosen to fit the distribution of case notification rates, and the knots outside the variable range were added for increased flexibility. For the splines of dates, three knots were placed at time points representing major changes in overall COVID-19 testing strategy: 12 October 2020, when the Gottsunda testing station was opened and centralized testing thus abandoned; 22 February 2021, when the testing of children ≥5 years was initiated; and 13 December 2021, when the testing strategy shifted from assisted PCR tests towards self-sampling PCR kits. Two additional knots were placed in the beginning (13 August 2020) and towards the end of the study period (15 January 2022) to improve statistical modelling.

**Difference-in-difference analysis comparing Gottsunda and Sävja**

The neighbourhood Gottsunda comprised the 7 postal code areas 75645, 75649, 75650, 75654, 75656, 75657, and 75658, and the neighbourhood Sävja the 2 postal code areas 75754 and 75755. For the difference-in-difference analysis we conducted the analysis on neighbourhood level instead of postal code level. Thus, the outcome here is daily number of tests per sex, age group and neighbourhood (either Sävja or Gottsunda). We used a Poisson model with regular robust standard errors and adjusted for the same covariates as the main model, except that we used a binary variable for Sävja/Gottsunda instead of Uppsala County/Uppsala City and that we used a binary intervention variable as exposure instead of CNI. Interaction terms were added in the model, whereby two-, three- and four-way interaction terms were added between all combinations of the variables intervention, sex, age category and date were fitted similarly to the main model.

**SUPPLEMENTARY TABLES**

**Supplementary Table 1.** Baseline postal code area characteristics in Uppsala County and Uppsala City, weighted for total population per postal code area. Uppsala County comprise postal codes beginning with 74 and 81, and Uppsala City beginning with 75. Values are presented as median (first and third quartiles), unless stated otherwise.

|  | Total | Uppsala County | Uppsala City |
| --- | --- | --- | --- |
| Postal code areas | N=350 | N=203 | N=147 |
| Care Need Index | 1.0  (0.8, 1.4) | 0.8  (0.7, 1.1) | 1.1  (0.8, 1.6) |
| Mean age, years | 40.6  (37.5, 43.2) | 41.6  (38.9, 43.8) | 39.7  (35.4, 42.4) |
| Proportion of women, % | 49.9  (48.3, 51.6) | 49.0  (47.6, 50.2) | 51.0  (49.3, 52.6) |
| Proportion of inhabitants <5 yearsꭞ, % | 5.6 (4.2, 6.6) | 5.8 (5.0, 6.8) | 5.1 (3.9, 6.3) |
| Proportion of inhabitants born in east- or south Europe (outside the European Union), Africa, Asia or South Americaꭞ, % | 6.2 (3.5, 14.4) | 3.9 (2.2, 6.6) | 10.5 (5.7, 19.8) |
| Proportion of inhabitants >65 years who reside in single-person householdsꭞ, % | 38.3  (29.7, 49.3) | 34.3  (28.4, 42.9) | 43.7  (31.6, 53.2) |
| Proportion of inhabitants who are single parents with children <18 yearsꭞ, % | 2.6 (2.0, 3.3) | 2.7 (2.2, 3.3) | 2.3 (1.8, 3.2) |
| Proportion of inhabitants >1 years who moved into area within the previous calendar yearꭞ, % | 8.2 (6.2, 11.3) | 6.8 (5.3, 8.5) | 10.0 (7.7, 13.5) |
| Proportion of inhabitants with compulsory education only^1^ꭞ, % | 13.1  (10.0, 20.0) | 12.9  (9.9, 17.9) | 13.3  (10.3, 22.3) |
| Proportion of inhabitants unemployed or enrolled in labour market measures^2^ꭞ, % | 7.3  (4.1, 10.6) | 9.5  (6.5, 11.3) | 4.5  (2.4, 8.9) |
| Distance to the nearest testing station^3^, km | 7.5 (3.3, 24.3) | 24.2 (11.6, 33.9) | 4.1 (2.6, 7.2) |

ꭞ Sociodemographic variable included in the composite measure Care Need Index.

^1^ In the population aged 25–64 years

^2^ In the population aged 16–64 years

^3^ Only applicable 24 June–11 October 2020

**Supplementary Table 2.** Baseline postal code area characteristics for the two Uppsala City neighbourhoods, Gottsunda and Sävja, weighted for total population per postal code area. Values are presented as median (first and third quartiles) unless stated otherwise.

|  | Gottsunda | Sävja |
| --- | --- | --- |
| Postal code areas | N=7 | N=2 |
| Care Need Index | 2.8 (2.1, 2.8) | 1.4 (1.4, 1.8) |
| Mean age, years | 34.9 (31.8, 38.2) | 40.0 (35.7, 40.0) |
| Proportion of women, % | 47.1 (46.7, 51.0) | 52.6 (50.5, 52.6) |
| Proportion of inhabitants <5 yearsꭞ, % | 8.5 (5.8, 9.6) | 6.0 (5.8, 6.0) |
| Proportion of inhabitants born in east- or south Europe (outside the European Union), Africa, Asia or South Americaꭞ, % | 48.5 (32.3, 49.3) | 19.4 (19.4, 28.1) |
| Proportion of inhabitants >65 years who reside in single-person householdsꭞ, % | 56.1 (52.9, 59.4) | 36.5 (36.5, 42.9) |
| Proportion of inhabitants who are single parents with children <18 yearsꭞ, % | 4.6 (3.8, 5.0) | 3.7 (3.7, 3.9) |
| Proportion of inhabitants >1 years who moved into area within the previous calendar yearꭞ, % | 9.3 (8.7, 12.2) | 8.8 (8.8, 9.3) |
| Proportion of inhabitants with compulsory education only^1^ꭞ, % | 43.7 (30.3, 49.1) | 21.4 (21.4, 25.9) |
| Proportion of inhabitants who are unemployed or enrolled in labour market measures^2^ꭞ, % | 18.5 (12.8, 21.4) | 9.0 (9.0, 13.6) |
| Distance to the nearest testing station^3^, km | 5.7 (1.8, 6.9) | 8.7 (8.7, 9.1) |

ꭞ Sociodemographic variable included in the composite measure Care Need Index.

^1^ In the population aged 25-64 years

^2^ In the population aged 16-64 years

^3^ Only applicable 24 June–11 October 2020

**Supplementary Table 3.** Main model. Highest and lowest test rate ratios (TRRs) with 95 confidence intervals (CIs) for postal code area Care Need Index (CNI) per sex and age groups in Uppsala County and Uppsala City across the three pandemic waves. This main model is adjusted for date, day of week of test, age group, sex, Uppsala County/Uppsala City, and daily case notification rates per age and sex group per 100 000 per postal code area.

|  | Second pandemic wave  12 November 2020–5 January 2021 | | | | Third pandemic wave  20 March 2020–5 May 2021 | | | | Fourth pandemic wave  2 January 2022–9 February 2022 | | | |
| --- | --- | --- | --- | --- | --- | --- | --- | --- | --- | --- | --- | --- |
|  | Uppsala City |  | Uppsala County |  | Uppsala City |  | Uppsala County |  | Uppsala City |  | Uppsala County |  |
|  | Highest TRR (95% CI) | Lowest TRR (95% CI) | Highest TRR (95% CI) | Lowest TRR (95% CI) | Highest TRR (95% CI) | Lowest TRR (95% CI) | Highest TRR (95% CI) | Lowest TRR (95% CI) | Highest TRR (95% CI) | Lowest TRR (95% CI) | Highest TRR (95% CI) | Lowest TRR (95% CI) |
| Women, ages 5-14 | **0.66 (0.59, 0.74)** | **0.61 (0.52, 0.72)** | **0.65 (0.56, 0.75)** | **0.63 (0.53, 0.75)** | **0.67 (0.61, 0.74)** | **0.63 (0.57, 0.70)** | **0.65 (0.57, 0.75)** | **0.63 (0.54, 0.73)** | **0.87 (0.81, 0.95)** | **0.72 (0.67, 0.77)** | **0.73 (0.62, 0.88)** | **0.65 (0.57, 0.75)** |
| Men, ages 5-14 | **0.62 (0.54, 0.70)** | **0.56 (0.47, 0.66)** | **0.72 (0.62, 0.85)** | **0.67 (0.54, 0.84)** | **0.65 (0.59, 0.72)** | **0.62 (0.57, 0.68)** | **0.73 (0.65, 0.82)** | **0.70 (0.62, 0.78)** | **0.88 (0.81, 0.95)** | **0.72 (0.66, 0.77)** | **0.79 (0.69, 0.90)** | **0.69 (0.61, 0.79)** |
| Women, ages 15-29 | **0.84 (0.79, 0.89)** | **0.81 (0.75, 0.88)** | 1.06 (0.96, 1.17) | 0.99 (0.91, 1.07) | **0.86 (0.82, 0.90)** | **0.84 (0.80, 0.89)** | **0.91 (0.84, 0.98)** | **0.89 (0.83, 0.96)** | 0.97 (0.90, 1.04) | **0.89 (0.83, 0.95)** | 0.96 (0.88, 1.03) | 0.95 (0.86, 1.04) |
| Men, ages 15-29 | **0.86 (0.81, 0.92)** | **0.84 (0.78, 0.91)** | 0.98 (0.90, 1.08) | 0.96 (0.89, 1.03) | **0.86 (0.80, 0.92)** | **0.83 (0.77, 0.89)** | 0.92 (0.85, 1.00) | **0.90 (0.83, 0.97)** | 1.00 (0.93, 1.07) | **0.90 (0.84, 0.96)** | 0.95 (0.83, 1.08) | 0.92 (0.83, 1.01) |
| Women, ages 30-49 | **0.74 (0.71, 0.77)** | **0.71 (0.68, 0.75)** | **0.79 (0.74, 0.85)** | **0.77 (0.71, 0.83)** | **0.78 (0.74, 0.83)** | **0.78 (0.74, 0.82)** | **0.82 (0.76, 0.88)** | **0.81 (0.75, 0.87)** | **0.89 (0.85, 0.94)** | **0.84 (0.80, 0.89)** | 0.93 (0.85, 1.02) | **0.86 (0.80, 0.92)** |
| Men, ages 30-49 | **0.82 (0.77, 0.87)** | **0.80 (0.75, 0.86)** | **0.87 (0.81, 0.93)** | **0.86 (0.80, 0.93)** | **0.84 (0.79, 0.89)** | **0.83 (0.78, 0.89)** | **0.87 (0.80, 0.95)** | **0.87 (0.79, 0.95)** | **0.93 (0.89, 0.97)** | **0.87 (0.84, 0.91)** | **0.85 (0.75, 0.97)** | **0.84 (0.75, 0.94)** |
| Women, ages 50-69 | **0.87 (0.82, 0.92)** | **0.84 (0.79, 0.90)** | 0.97 (0.90, 1.05) | 0.93 (0.85, 1.02) | **0.90 (0.86, 0.95)** | **0.90 (0.85, 0.95)** | 1.00 (0.92, 1.09) | 0.98 (0.90, 1.07) | **0.94 (0.89, 0.99)** | **0.91 (0.87, 0.94)** | 1.02 (0.93, 1.13) | 0.95 (0.88, 1.03) |
| Men, ages 50-69 | **0.83 (0.80, 0.87)** | **0.81 (0.77, 0.86)** | 0.97 (0.87, 1.07) | 0.95 (0.88, 1.04) | **0.87 (0.82, 0.93)** | **0.86 (0.81, 0.92)** | 0.95 (0.85, 1.06) | 0.92 (0.82, 1.03) | **0.93 (0.87, 0.99)** | **0.87 (0.82, 0.91)** | 0.99 (0.88, 1.12) | 0.91 (0.82, 1.03) |
| Women, ages 70+ | **1.19 (1.04, 1.37)** | **1.18 (1.04, 1.35)** | **1.40 (1.11, 1.77)** | **1.36 (1.11, 1.67)** | 1.08 (0.93, 1.24) | 0.98 (0.86, 1.13) | **1.19 (1.01, 1.40)** | 1.09 (0.94, 1.27) | 1.17 (0.89, 1.53) | 1.00 (0.81, 1.24) | 1.27 (0.98, 1.64) | 1.14 (0.93, 1.40) |
| Men, ages 70+ | **1.15 (1.01, 1.31)** | 1.11 (0.95, 1.30) | **1.28 (1.08, 1.52)** | **1.25 (1.09, 1.44)** | 1.15 (0.97, 1.35) | 1.08 (0.93, 1.26) | 1.14 (0.99, 1.31) | 1.04 (0.91, 1.20) | 0.94 (0.71, 1.24) | 0.87 (0.71, 1.06) | **1.25 (1.00, 1.56)** | 1.08 (0.89, 1.32) |

**Supplementary Table 4.** Sensitivity analysis. Highest and lowest test rate ratios (TRRs) with 95 confidence intervals (CIs) for Care Need Index (CNI) in sex and age groups in Uppsala County and Uppsala City across the three pandemic waves. This sensitivity analysis model is adjusted for date, day of week of test, age group, sex, Uppsala County/Uppsala City, and daily hospital admissions per 100 000 per postal code area.

|  | Second pandemic wave  November 12, 2020-January 5, 2021 | | | | Third pandemic wave  March 20, 2020 to May 5, 2021 | | | | Fourth pandemic wave  January 2, 2022 to February 9, 2022 | | | |
| --- | --- | --- | --- | --- | --- | --- | --- | --- | --- | --- | --- | --- |
|  | Uppsala city |  | Uppsala county |  | Uppsala city |  | Uppsala county |  | Uppsala city |  | Uppsala county |  |
|  | Highest TRR (95% CI) | Lowest TRR (95% CI) | Highest TRR (95% CI) | Lowest TRR (95% CI) | Highest TRR (95% CI) | Lowest TRR (95% CI) | Highest TRR (95% CI) | Lowest TRR (95% CI) | Highest TRR (95% CI) | Lowest TRR (95% CI) | Highest TRR (95% CI) | Lowest TRR (95% CI) |
| Women, ages 5-14 | **0.64 (0.57, 0.72)** | **0.60 (0.51, 0.72)** | **0.63 (0.52, 0.77)** | **0.63 (0.54, 0.74)** | **0.64 (0.57, 0.71)** | **0.61 (0.54, 0.68)** | **0.62 (0.53, 0.72)** | **0.60 (0.52, 0.70)** | **0.75 (0.66, 0.86)** | **0.65 (0.59, 0.72)** | **0.63 (0.50, 0.79)** | **0.60 (0.51, 0.70)** |
| Men, ages 5-14 | **0.60 (0.52, 0.68)** | **0.54 (0.45, 0.65)** | **0.71 (0.59, 0.84)** | **0.67 (0.53, 0.86)** | **0.63 (0.57, 0.71)** | **0.61 (0.55, 0.67)** | **0.70 (0.61, 0.79)** | **0.67 (0.59, 0.76)** | **0.77 (0.70, 0.86)** | **0.65 (0.60, 0.71)** | **0.64 (0.53, 0.78)** | **0.61 (0.51, 0.72)** |
| Women, ages 15-29 | **0.83 (0.78, 0.88)** | **0.80 (0.74, 0.87)** | 1.07 (0.97, 1.19) | 1.00 (0.92, 1.08) | **0.85 (0.80, 0.90)** | **0.83 (0.78, 0.89)** | **0.91 (0.84, 0.99)** | **0.89 (0.82, 0.97)** | 1.02 (0.91, 1.14) | 0.92 (0.84, 1.00) | 0.95 (0.85, 1.06) | 0.93 (0.80, 1.09) |
| Men, ages 15-29 | **0.84 (0.79, 0.90)** | **0.82 (0.76, 0.90)** | 0.99 (0.90, 1.10) | 0.96 (0.88, 1.04) | **0.84 (0.78, 0.91)** | **0.82 (0.76, 0.89)** | **0.90 (0.82, 0.99)** | **0.88 (0.81, 0.96)** | 0.99 (0.89, 1.11) | **0.90 (0.82, 0.99)** | **0.88 (0.78, 0.98)** | 0.88 (0.74, 1.04) |
| Women, ages 30-49 | **0.75 (0.71, 0.79)** | **0.72 (0.68, 0.76)** | **0.79 (0.73, 0.86)** | **0.78 (0.71, 0.85)** | **0.79 (0.74, 0.84)** | **0.78 (0.74, 0.83)** | **0.81 (0.76, 0.88)** | **0.81 (0.75, 0.87)** | 0.95 (0.87, 1.03) | **0.86 (0.80, 0.94)** | **0.81 (0.70, 0.95)** | **0.80 (0.71, 0.89)** |
| Men, ages 30-49 | **0.83 (0.78, 0.88)** | **0.81 (0.75, 0.87)** | **0.88 (0.81, 0.95)** | **0.86 (0.79, 0.94)** | **0.85 (0.80, 0.91)** | **0.84 (0.78, 0.90)** | **0.89 (0.81, 0.98)** | **0.88 (0.80, 0.97)** | 0.97 (0.91, 1.05) | **0.89 (0.84, 0.95)** | 0.85 (0.71, 1.01) | **0.83 (0.72, 0.96)** |
| Women, ages 50-69 | **0.90 (0.85, 0.96)** | **0.87 (0.81, 0.93)** | 0.98 (0.90, 1.06) | 0.94 (0.85, 1.04) | **0.94 (0.89, 0.99)** | **0.92 (0.87, 0.97)** | 1.00 (0.91, 1.10) | 0.98 (0.89, 1.07) | 1.07 (0.97, 1.18) | 0.98 (0.92, 1.04) | 1.03 (0.90, 1.18) | 0.96 (0.87, 1.06) |
| Men, ages 50-69 | **0.87 (0.82, 0.92)** | **0.84 (0.79, 0.90)** | 0.97 (0.87, 1.09) | 0.96 (0.87, 1.06) | **0.89 (0.83, 0.96)** | **0.87 (0.81, 0.94)** | 0.96 (0.84, 1.09) | 0.93 (0.82, 1.05) | 0.99 (0.90, 1.10) | **0.90 (0.84, 0.97)** | 1.02 (0.87, 1.19) | 0.93 (0.81, 1.06) |
| Women, ages 70+ | **1.20 (1.04, 1.38)** | **1.18 (1.01, 1.38)** | **1.42 (1.10, 1.84)** | **1.37 (1.10, 1.70)** | 1.10 (0.95, 1.28) | 1.00 (0.87, 1.16) | **1.18 (1.00, 1.39)** | 1.09 (0.93, 1.27) | 1.20 (0.89, 1.64) | 1.01 (0.81, 1.27) | 1.24 (0.95, 1.61) | 1.13 (0.91, 1.39) |
| Men, ages 70+ | **1.18 (1.02, 1.35)** | 1.13 (0.96, 1.32) | **1.31 (1.08, 1.58)** | **1.27 (1.09, 1.47)** | 1.17 (0.98, 1.40) | 1.10 (0.93, 1.29) | 1.14 (0.98, 1.31) | 1.04 (0.90, 1.19) | 0.96 (0.70, 1.32) | 0.88 (0.71, 1.10) | **1.29 (1.01, 1.66)** | 1.10 (0.88, 1.36) |

**Supplementary Table 5.** Highest and lowest test rate ratios (TRRs) with 95 confidence intervals (CIs) for distance to nearest main testing station (in kilometres) presented per sex and age groups in Uppsala County and Uppsala City for the earlier part of the study period (24 June 2020–11 October 2020). The model is adjusted for Care Need Index (CNI), date, day of week of test, age group, sex, Uppsala County/Uppsala City, and daily case notification rates per age and sex group per 100 000 per postal code area.

|  | Uppsala County |  | Uppsala City |  |
| --- | --- | --- | --- | --- |
|  | Highest TRR (95% CI) | Lowest TRR (95% CI) | Highest TRR (95% CI) | Lowest TRR (95% CI) |
| Women, ages 5-14 | **0.980 (0.968, 0.992)** | **0.973 (0.953, 0.993)** | **0.960 (0.923, 0.998)** | **0.943 (0.922, 0.964)** |
| Men, ages 5-14 | **0.985 (0.975, 0.995)** | **0.966 (0.949, 0.983)** | **0.960 (0.924, 0.997)** | **0.950 (0.916, 0.984)** |
| Women, ages 15-29 | **0.990 (0.983, 0.996)** | **0.980 (0.970, 0.990)** | 0.982 (0.963, 1.000) | **0.969 (0.949, 0.990)** |
| Men, ages 15-29 | **0.986 (0.979, 0.994)** | **0.984 (0.973, 0.995)** | 0.988 (0.970, 1.005) | **0.963 (0.937, 0.990)** |
| Women, ages 30-49 | 0.995 (0.991, 1.000) | **0.984 (0.977, 0.990)** | 1.001 (0.990, 1.011) | 0.997 (0.984, 1.010) |
| Men, ages 30-49 | 0.995 (0.989, 1.000) | **0.980 (0.971, 0.989)** | 0.991 (0.977, 1.005) | **0.973 (0.958, 0.987)** |
| Women, ages 50-69 | **0.994 (0.989, 0.998)** | **0.984 (0.978, 0.990)** | 0.998 (0.980, 1.016) | 0.983 (0.967, 1.000) |
| Men, ages 50-69 | 0.996 (0.986, 1.007) | **0.988 (0.981, 0.996)** | 0.992 (0.970, 1.014) | **0.979 (0.961, 0.998)** |
| Women, ages 70+ | 1.006 (0.988, 1.024) | 0.994 (0.980, 1.008) | 1.001 (0.955, 1.049) | 0.934 (0.867, 1.007) |
| Men, ages 70+ | 1.006 (0.986, 1.026) | 0.984 (0.968, 1.001) | 0.984 (0.925, 1.046) | 0.966 (0.921, 1.013) |

**SUPPLEMENTARY FIGURES**

**Supplementary Figure 1.** Placements of the four stations for patient-initiated COVID-19 PCR testing initially set up across Uppsala County in June 2020 - in the north, east, and northwest part of the county, and in the centre of Uppsala City.

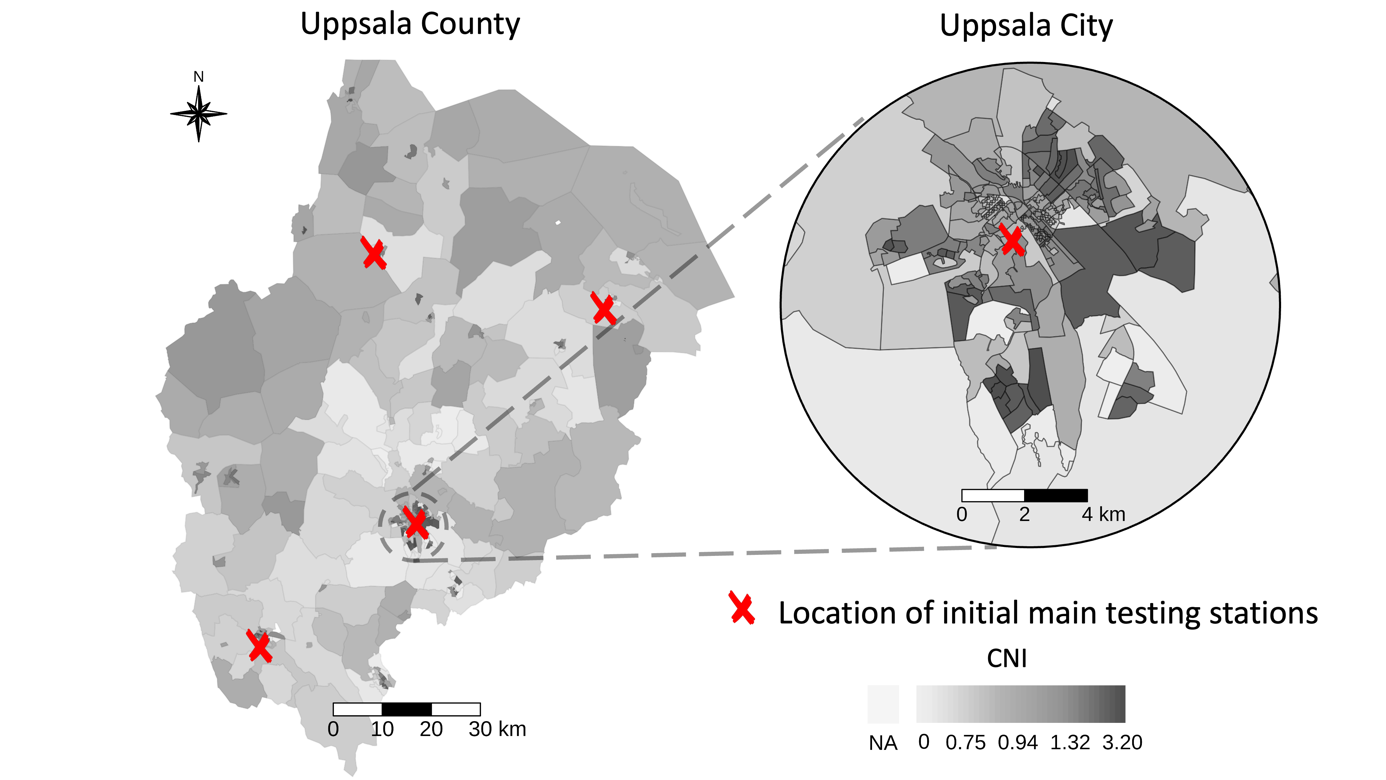

**Supplementary Figure 2.** The frequency distribution of postal code areas per Care Need Index (CNI) in Uppsala County (n=203) and Uppsala City (n=147). Higher CNI indicates higher primary health care burden.

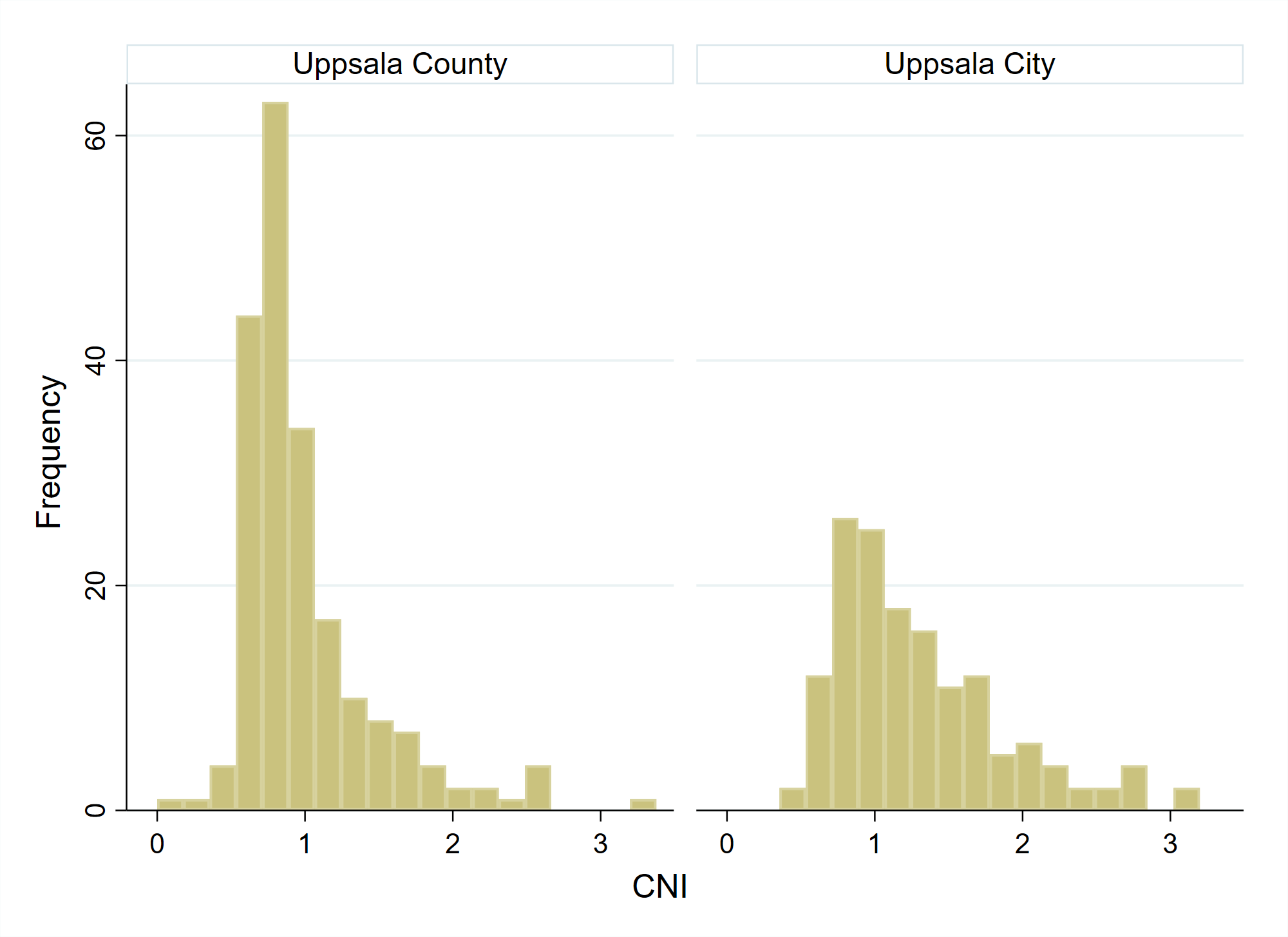

**Supplementary Figure 3.** Population-weighted Pearson correlation matrices for the composite measure Care Need Index (CNI), proportion of women (%), the seven sociodemographic variables included in CNI*, and distance to nearest main testing station (only applicable 24 June to October 11, 2020) in Uppsala County (left) and Uppsala City (right).

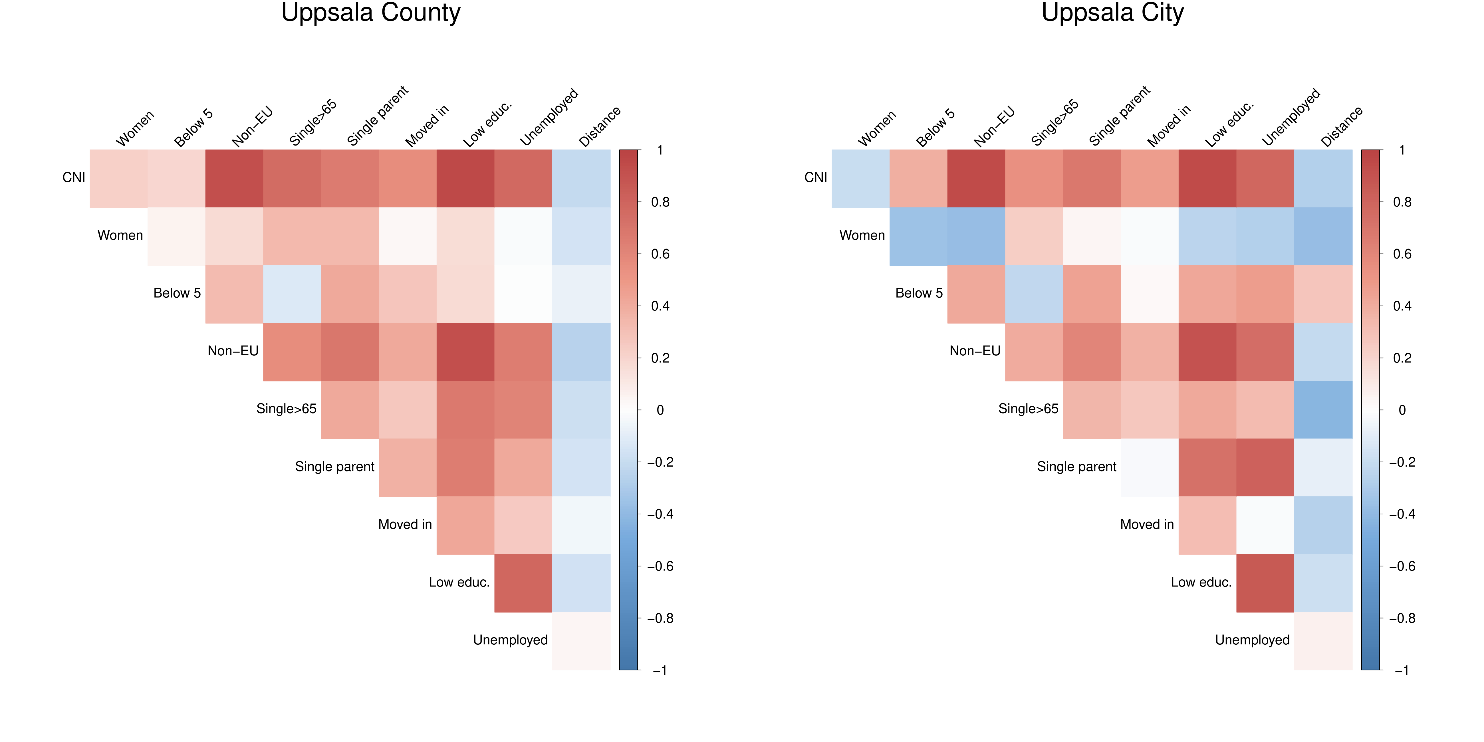

*****Proportion of inhabitants in a postal code area who were 1) <5 years, 2) born in East Europe (outside the European Union), Asia, Africa, or South America, 3) >65 years and reside in single-person households, 4) single parents with children <18 years, 5) >1 years and have moved into the postal code area within the previous calendar year, 6) 25–64 years with low educational attainment (≤9 years of schooling, equivalent to compulsory education only in Sweden), 7) 16–64 years and unemployed or enrolled in a labour market programme.

**Supplementary Figure 4.** 7-day rolling averages of daily number of COVID-19 PCR tests, positive tests and hospital admissions per 100 000 inhabitants across the study period (24 June 2020–9 February 2022) for inhabitants residing postal code areas in Uppsala County or Uppsala City.

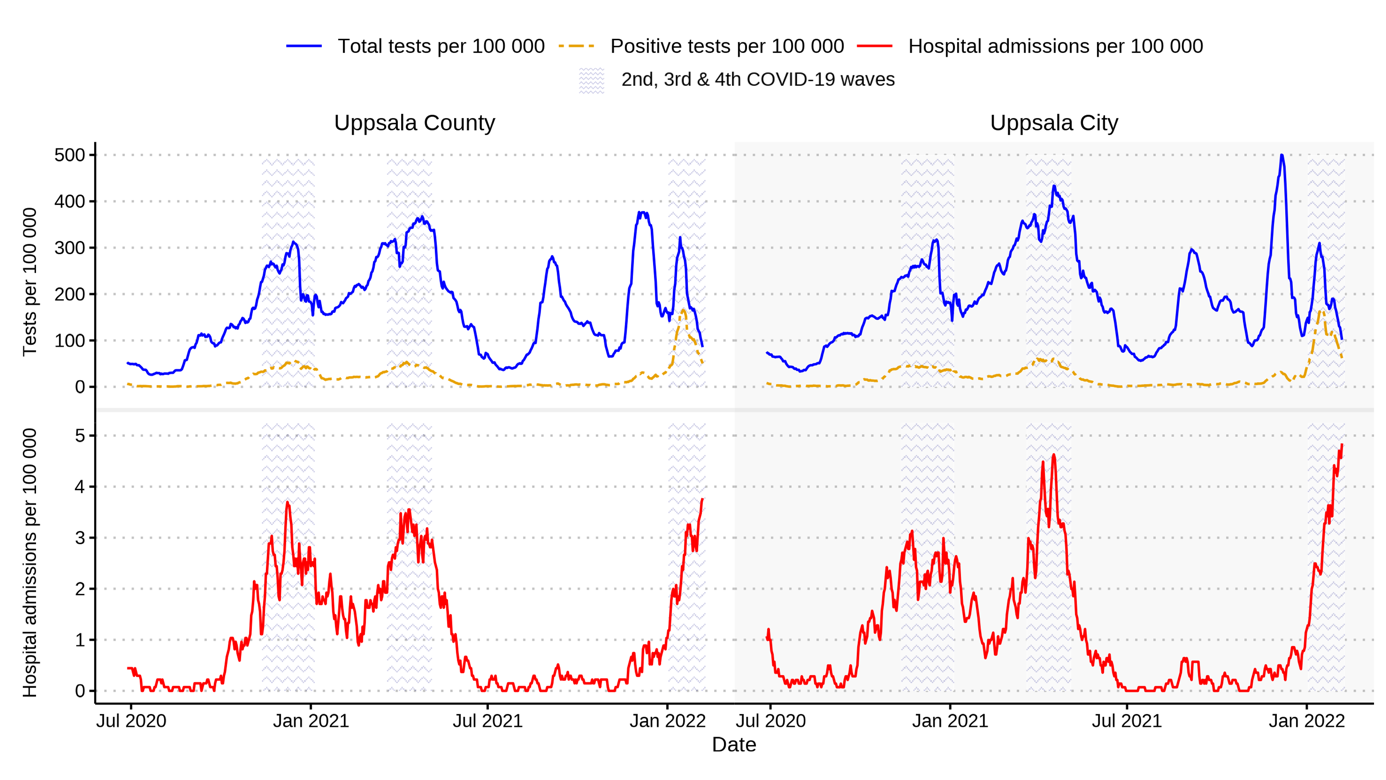

**Supplementary Figure 5.** 7-day rolling averages of daily number of COVID-19 PCR tests and positive tests per sex and age group per 100 000 inhabitants, across the study period (24 June 2020–9 February 2022) for inhabitants residing in a postal code area in Uppsala County (a) or Uppsala City (b).

a)

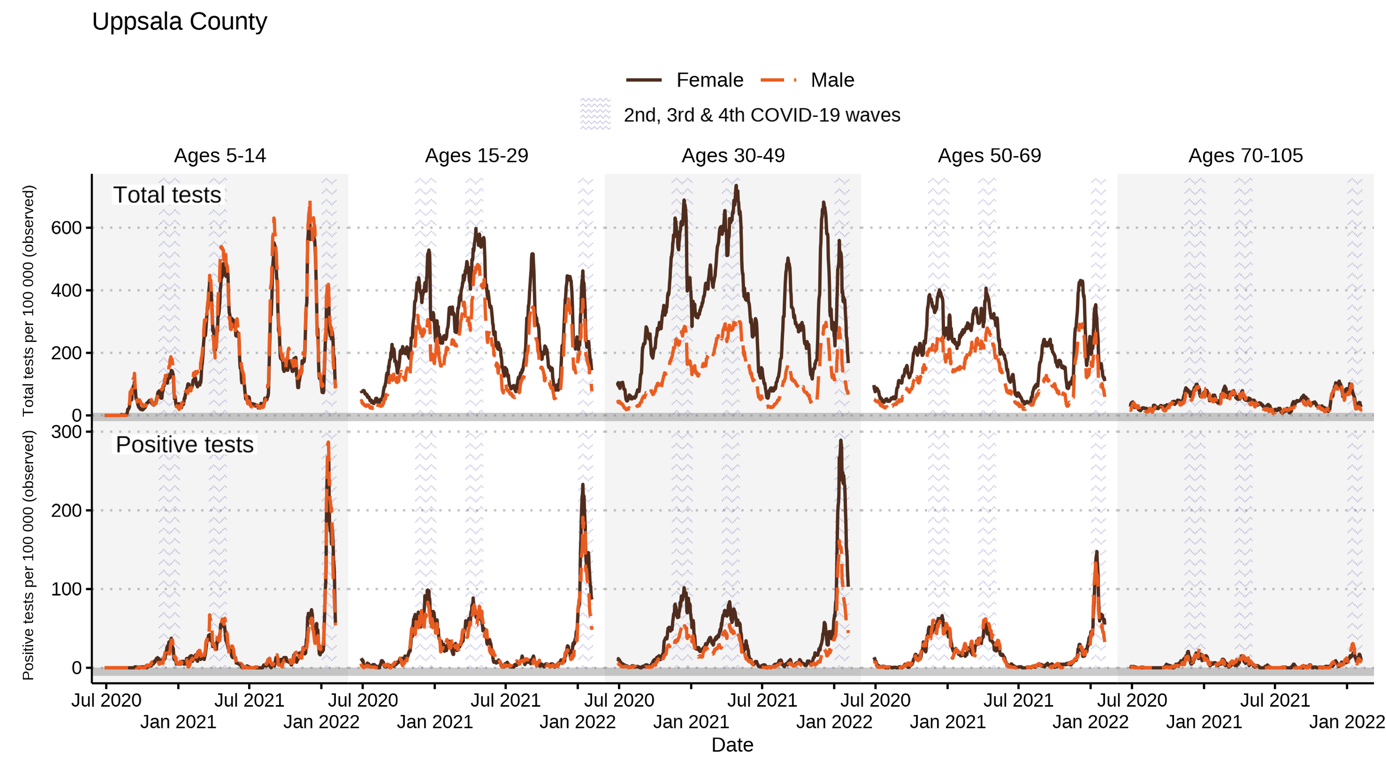

b)

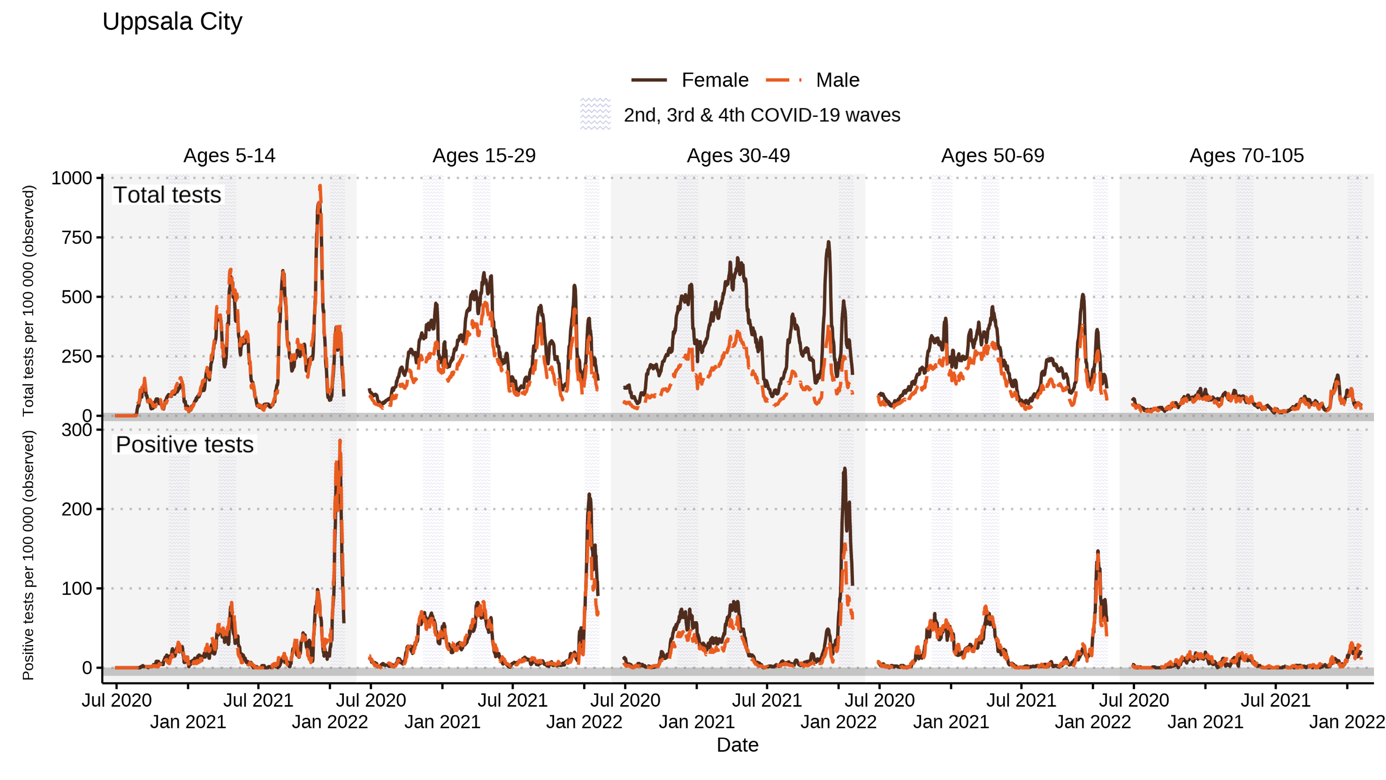

**Supplementary Figure 6.** Population-weighted cumulative COVID-19 vaccination coverage (defined as ≥2 doses) in inhabitants 15-105 years in Uppsala County (left) and Uppsala City (right), by quartiles of Care Need Index (CNI; Q1 represents lowest CNI quartile). Only vaccinations administrated at healthcare units within Uppsala County Council are included in these data.

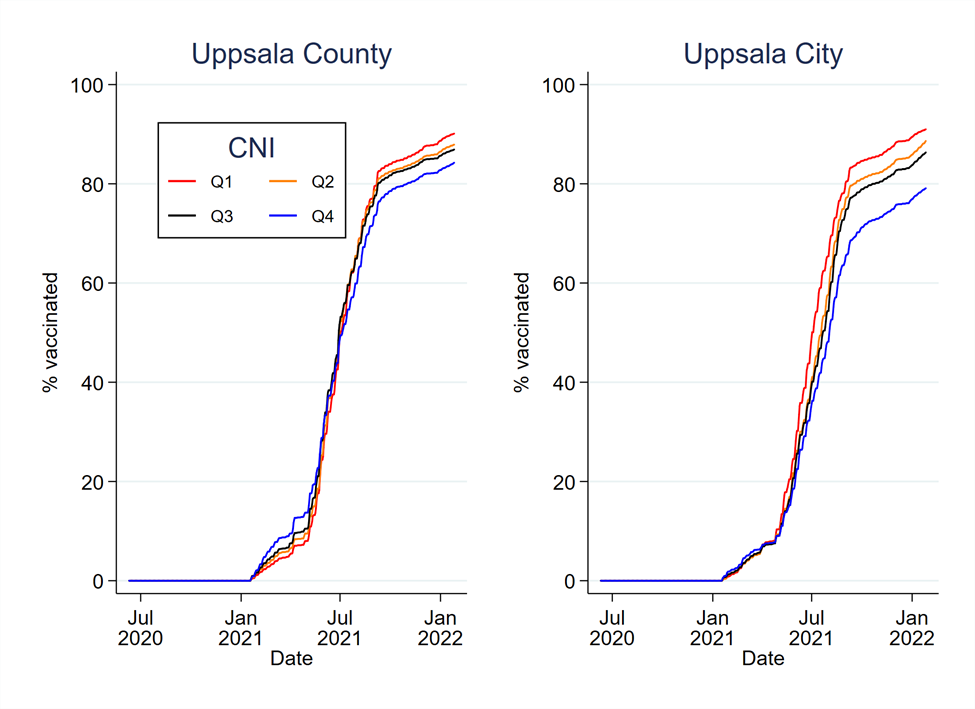

**Supplementary Figure 7.** Sensitivity analysis. Model-based testing rates for COVID-19 per sex and age group per 100 000 inhabitants in Uppsala County (not including Uppsala City) (a) and Uppsala City (b) across the study period (24 June 2020–9 February 2022), presented for the 10^th^ and 90^th^ postal code area for the Care Need Index (CNI) percentiles. Models are adjusted for number of daily COVID-19 hospital admissions per 100 000 inhabitants.

a)

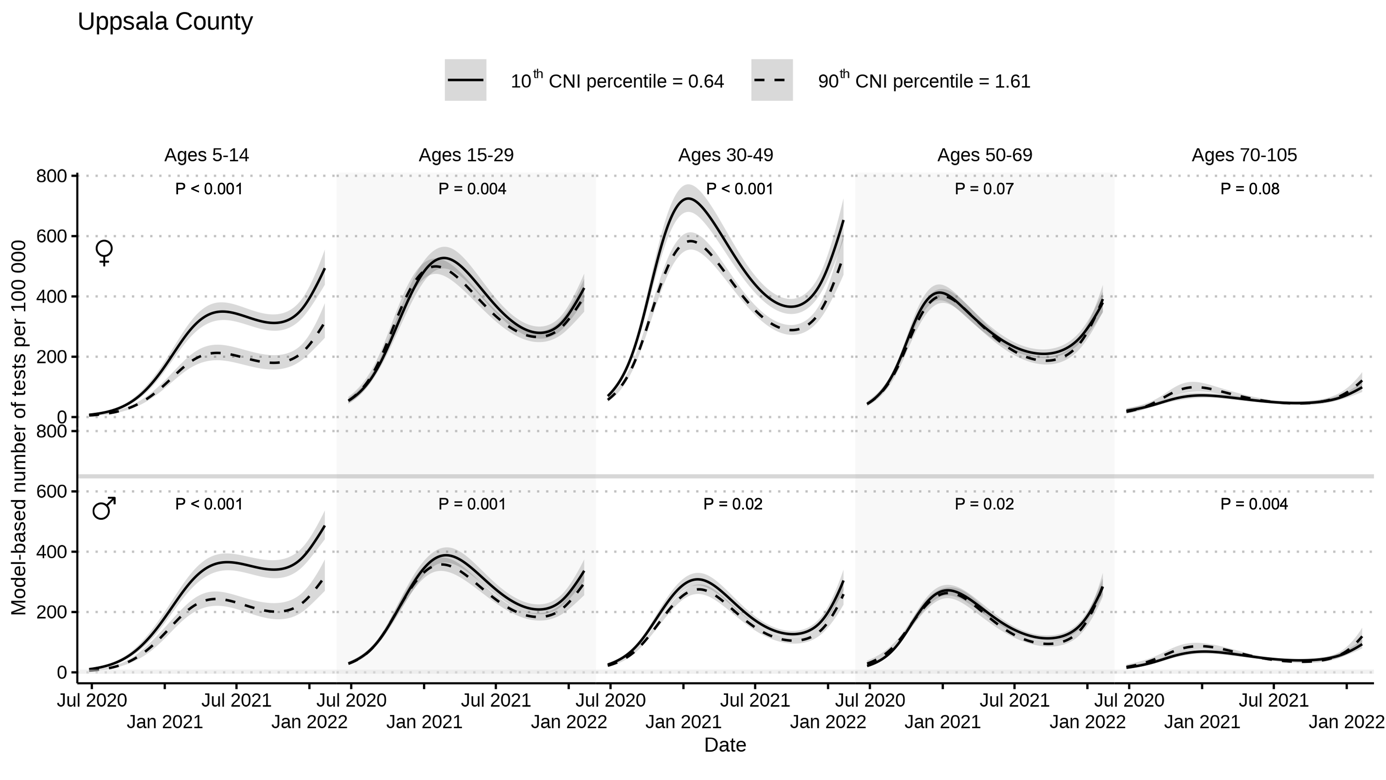

b)

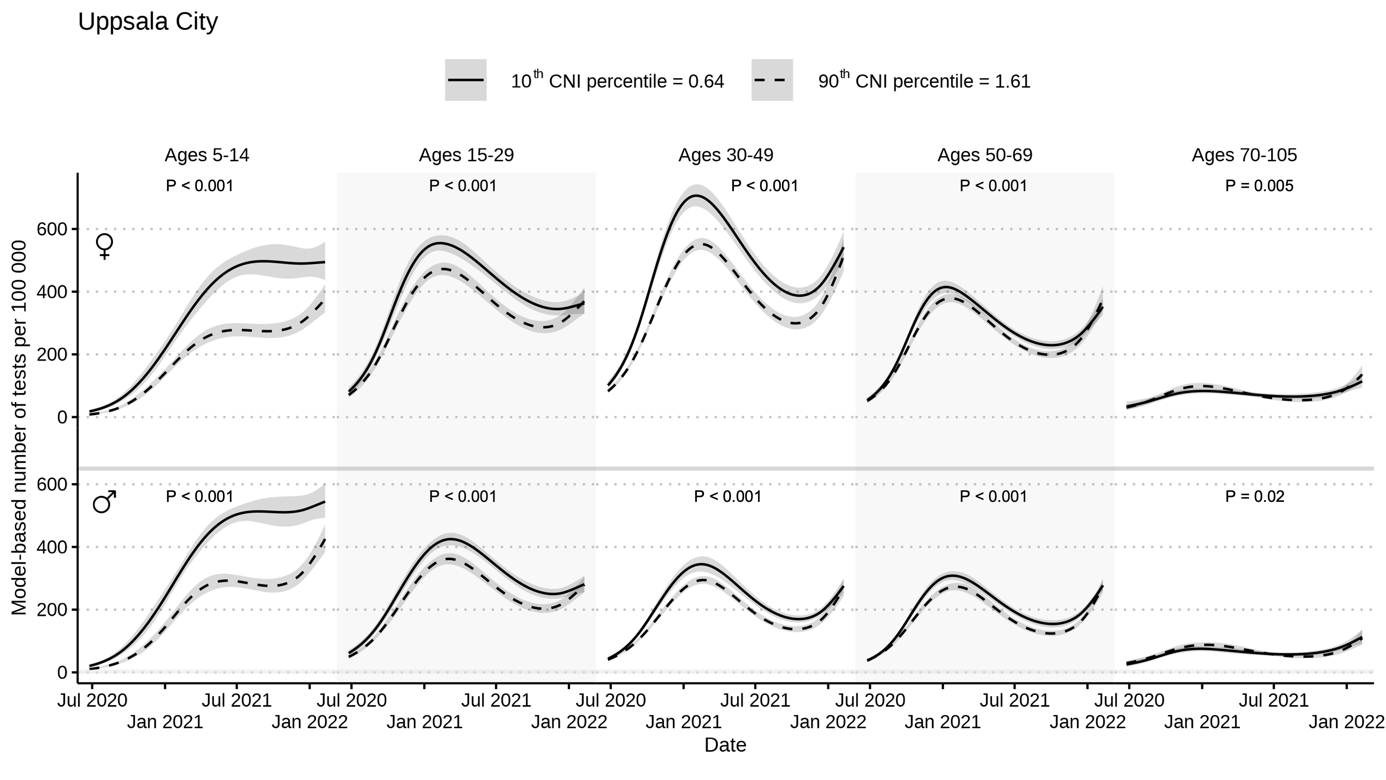

**Supplementary Figure 8.** Model based testing rates for COVID-19 per sex and age group per 100 000 inhabitants in Uppsala County (not including Uppsala City) (a) and Uppsala City (b) across the early part of the study period (24 June 24–11 October 2020), presented for the 10^th^ and 90^th^ postal code area for the distance to nearest testing station (in kilometres).

a)

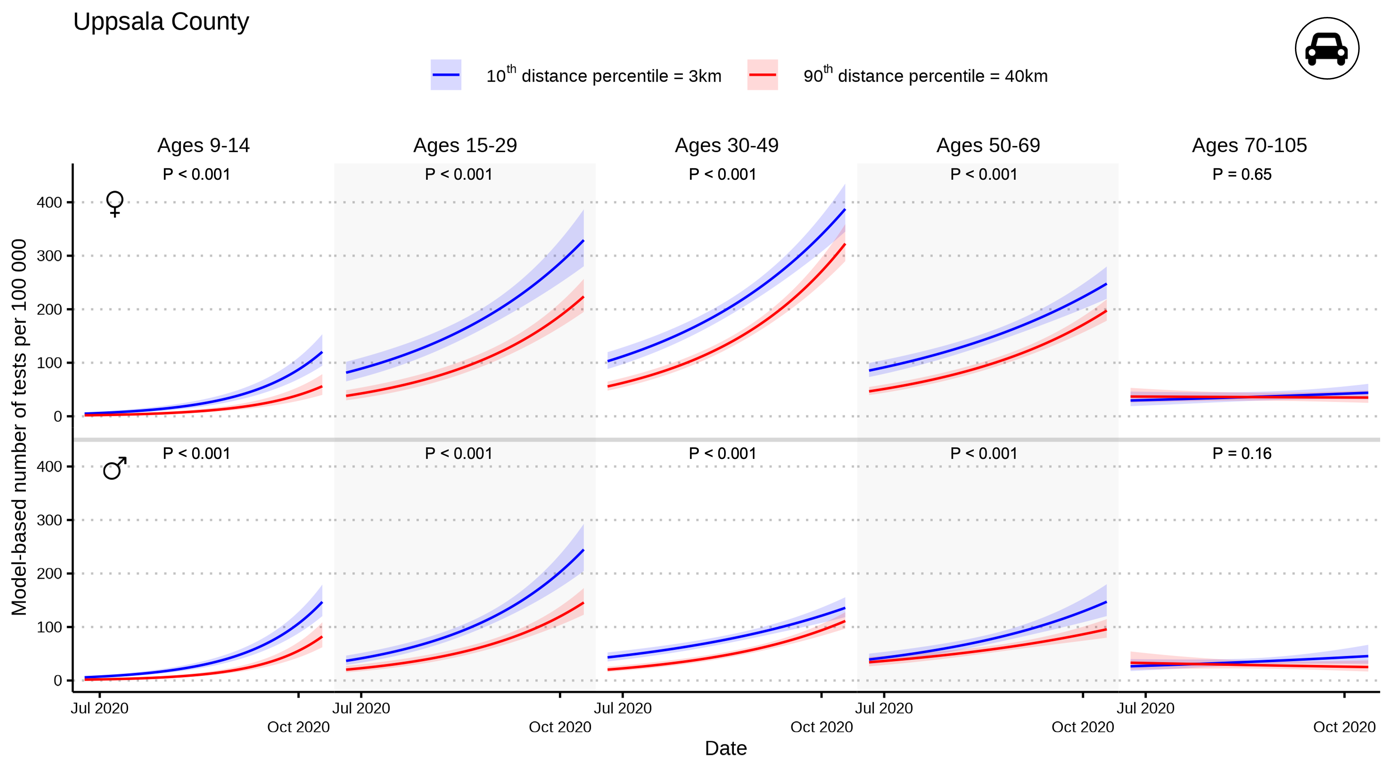

b)

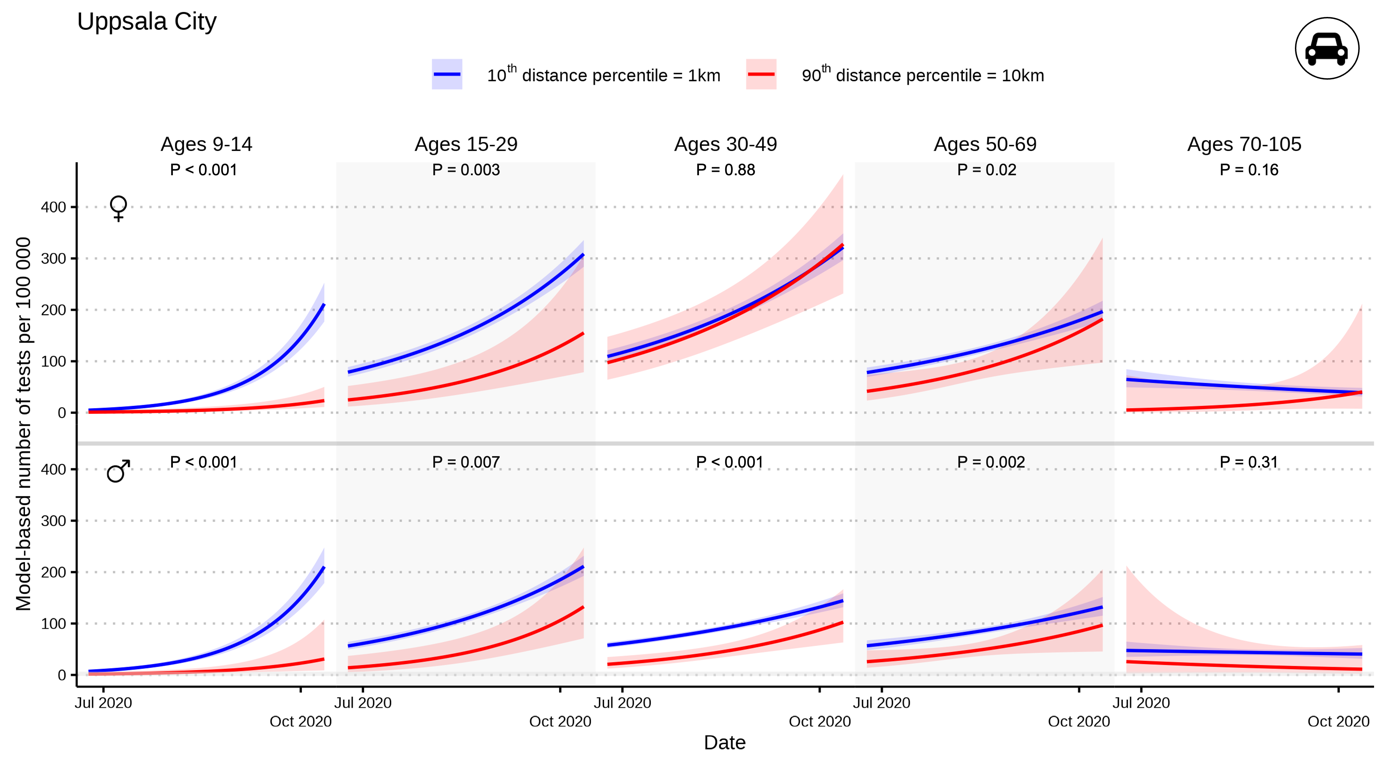
